# Integrating voluntary family planning and cervical cancer screening and preventative treatment: lessons from the MULHER study in Mozambique

**DOI:** 10.64898/2026.11.02.26366931

**Authors:** Marcos Chissano, Paul Bouanchaud, Edson Chivambo, Yolanda Chongo, Hannah Hoover, Guilherima Tivir, Pontremoli Salcedo Mila, Kathleen M Schmeler, Eva Lathrop

## Abstract

**Introduction:** Mozambique faces a significant burden of cervical cancer, but screening rates are currently low. While voluntary family planning (vFP) services are more widely accessible, unmet need for contraception remains at 22%. Integrating vFP and cervical cancer screening and preventive therapy (CCS&PT) services presents an opportunity to address these issues by improving service efficiency and reaching more women. This paper examines the uptake, feasibility, and experiences of integrating vFP services into CCS&PT services in Mozambique.

**Methods:** This mixed-methods operational research study was conducted in four public health facilities in Mozambique. Quantitative data were collected from January 2020 to October 2022 as part of the MULHER study and included program monitoring data from clinical encounters (N=9,014) and client exit interviews (N=504). Qualitative data was gathered through in-depth interviews (IDIs) with 60 clients and 15 providers in two phases, in October 2021 and November 2022.

**Results:** Out of 9,014 women eligible for HPV screening, 88% received both HPV screening and vFP counselling. 21% of these women took up a modern method of FP, 20% of whom were new users. Multivariable analysis showed that the likelihood of receiving integrated services was significantly associated with a range of factors including parity, age and HIV status. Qualitative data revealed that both clients and providers generally perceived the integrated model as beneficial. Clients highlighted the convenience, enhanced privacy, reduced travel costs, and opportunity to learn about new services. Providers emphasized similar benefits alongside the potential for increased demand through peer referrals.

**Conclusions:** Integrating vFP into CCS&PT services in Mozambique has the potential to increase vFP uptake, including among new users. The integrated model proved feasible and acceptable to both women and providers. The findings support the scaling up of this model to improve access to vFP and CCS&PT services and enhance women’s sexual and reproductive health.

**Teaser key message:** Integrating voluntary family planning into cervical cancer screening service provision in Mozambique results in high levels of satisfaction among patients and providers and appears to make FP methods available to new users.

**Key Messages:**

1. The study shows high levels of uptake of integrated vFP services among women enrolled in cervical screening and preventive therapy (CCS&PT) in the four public health facilities in Maputo and Gaza provinces, with almost 9 in 10 women who were enrolled for HPV screening also receiving vFP counselling on the day of their visit.
2. Offering vFP services to women who attend CCS&PT services in Mozambique is feasible and presents an opportunity for increased access to women with unmet needs; one in five of the women taking up a contraceptive method were new users.
3. Clients who accessed integrated services expressed a high degree of satisfaction with the convenience, enhanced privacy, reduced travel costs and opportunity to learn about new services that this model presents.

## Introduction

Mozambique has one of the highest burdens of cervical cancer in the world, with an incidence of 42.8 and mortality rate of 35.7 per 100,000 women (Ferlay et al., 2015; Ferlay et al., 2024). The prevalence of HIV, a significant risk factor for cervical cancer (World Health Organization, 2018), is 15% nationally among women over 15 years (INSIDA, 2021) and >20% in some urban regions. Mozambique has a shortage of medical providers with only three doctors per 100,000 people, only 17 pathologists in the national health system (1 per 2 million) and seven medical oncologists (1 per 9 million) in the entire country (Ismail et al., 2021). Currently in Mozambique, it is estimated that fewer than 5% of eligible women are undergoing cervical cancer screening, while many women testing positive are lost to follow- up and not receiving the necessary evaluation and procedures to diagnose and treat precancerous cervical lesions (Brandão et al., 2018). Most cases of cervical cancer in Mozambique are diagnosed at an advanced stage (Tulsidás et al., 2024). Voluntary family planning (vFP) services meanwhile are more widely available, with a modern contraceptive prevalence rate (mCPR) among all women of 32% in 2022 (Track20, n.d.), but gaps in service provision and quality (Pires et al., 2022) persist. Unmet need for contraception in Mozambique is estimated at 22.4% (Track20, n.d.), and in 2022 there were an estimated 300,000 unintended pregnancies in the country (Track20, n.d.).

Integration is a high impact practice and has largely been implemented through combining vFP with HIV, immunization, or Antenatal Care/Postnatal Care (ANC/PNC) interventions. In the context of recent contractions in available external funding for vFP and HIV (Stover et al., 2025), there is increased pressure on government to find ways to deliver services more cost efficiently, making integrated approaches with fewer contacts a practical option. The integration of vFP and Cervical Cancer Screening and Preventive Therapy (CCS&PT) is less commonly reported in the literature. There is a nascent body of literature showing the feasibility and cost-effectiveness of integrating cervical cancer screening and preventive treatment (CCS&PT) with other services, including HIV and or vFP, with evidence from several countries in sub-Saharan Africa, suggesting CCS integration models can increase the uptake of vFP and HIV services for women (White et al., 2017). Here we define integration of vFP and CCS&PT as clients having same-visit access to both quality services, provided by one provider or multiple providers working as a team. Several models to integration exist. Recent evidence from a Malawi cluster randomized trial has shown that an integrated model of CCS&PT and vFP at both community and clinic level led to an increased uptake of both services by women, when compared to a clinic only model (Tang et al., 2023). To our knowledge no evidence has been published to date that examines the results of CCS&PT and vFP integration in the Mozambique context.

Mozambique’s Strategic Health Plan for 2014 to 2019, extending to at least 2024 (MISAU, 2022) established the promotion of universal health care, with a focus on Primary Health Care, while the most recent 2023-2030 National Family Planning strategy calls specifically for the expansion of integrated vFP services (Direcção Nacional de Saúde Pública, 2023). This means providing integrated and continuous services that are accessible, locally relevant and acceptable, with appropriate technology, participation and community ownership. The program includes the integration of cervical cancer screening into sexual and reproductive health (SRH) programs. In 2009, the Mozambique Ministry of Health initiated the program for prevention of cervical and breast cancer, having already established basic cervical cancer screening services in 75 health facilities in country. Despite policy level efforts to promote both CCS&PT and service integration more widely, anecdotal evidence suggests implementation of this policy is patchy.

This manuscript presents evidence from operational research conducted as part of the Partnership for Enhanced Engagement Research (PEER) project in Mozambique and MULHER Study, in which vFP services were integrated into CCS&PT services in four public health facilities as part of a larger clinical trial of CCS&PT diagnostics and treatments (Salcedo et al., 2023; Lathrop et al., 2023; Batman et al., 2023).

We examine uptake of FP counselling and services among CCS&PT clients, the feasibility of implementing this integrated model in Mozambique, and aspects of client and provider experience with this form of service provision. We define integrated care here as vFP and CCS&PT services delivered by a single provider or a coordinated team, ensuring clients receive quality care for both services during the same visit. We present recommendations for policy makers and programmers aiming to develop, refine or scale-up similar integrated services.

## Methods

This section outlines the quantitative and qualitative data collection and analysis approaches taken in this mixed-methods operational research study conducted in four public health facilities in two provinces of Mozambique where the PEER program’s CCS&PT services were delivered. The study methods discussed here pertain specifically to the integrated vFP and CCS&PT services but were a part of a larger suite of research activities that are reported elsewhere (Salcedo et al., 2023). The components of the present study were designed to allow for triangulation of findings, with the objective of gaining a comprehensive view of the facilitators of and barriers to integration, and experiences of those who engaged with the project offering integrated cervical cancer screening and family planning provision.

The Partnership for Enhanced Engagement Research (PEER) was a collaborative project between Eduardo Mondlane University Faculty of Medicine (UEM), The University of Texas MD Anderson Cancer Center (MD Anderson), the United States Agency for International Development and National Academy of Science, Engineering and Medicine (USAID/NASEM) and Population Services International (PSI), to generate evidence to inform national and global efforts to scale-up effective and efficient cervical cancer screening and treatment programs. The consortium aimed to evaluate the impact of integration of cervical cancer prevention into existing vFP programs in Mozambique, while testing the feasibility and acceptability of innovative point of care molecular diagnostic tests for detection of high-risk human papillomavirus (hrHPV), and to strengthen capacity to implement these CCS&PT interventions more efficiently and effectively at scale. The PEER project’s cervical cancer screening services with hrHPV test were incorporated into pre- existing clinic services, vFP services, and were integrated into clinic staff procedures.

The quantitative dataset presented here comprises program monitoring data from clinical encounters (N=9,014) and client exit interview data (N=504). The program monitoring data were collected routinely during the implementation of the MULHER study (January 2020 to January 2023). Women aged 30-49 were prospectively enrolled from January 2020 to October 2022 in Maputo City and Gaza province, Mozambique. These client-level data were recorded in pen and paper client records by trained providers working in the PEER project. They contained variables describing client characteristics, HPV sample type (self-sampling or provider collected sample), vFP counselling and FP product use, alongside information on hrHPV test results and subsequent follow-up. All data was entered in REDCap (Harris et al., 2009), a web application for building and managing online surveys and databases.

These client-level data were used to estimate monthly vFP and CCS&PT services provision numbers at each intervention site at baseline, prior to integration roll-out (based on a three- month average), while subsequent client numbers following implementation of service integration were estimated monthly.

The qualitative component consisted of in-depth interviews (IDIs) (N=75), to evaluate the experience with, and perceptions of the integrated model CCS&PT and vFP services, incorporating new screening and treatment technologies. Each interview followed a structured guide, covering themes including views of integrated services, preferences for self- collection versus provider collection for CCS, challenges experienced within health facilities, and perceptions of barriers and facilitators to screening access.

Data collection occurred in two phases: the first wave (N clients=20) took place in October 2021, followed by second wave (N clients =40; N providers = 15) in November 2022. Clients were recruited from waiting rooms, provided with a study overview, and enrolled upon giving written consent. Data was gathered from two health facilities in Maputo and two in Gaza province, and thematic analysis was performed using Dedoose (Dedoose, 2023), a qualitative analysis software package.

## Research ethics and data availability

All research participants gave their informed consent to take part in the study. This study was determined to be human subjects research and ethics approval for the study was granted by the Moçambique Comité Nacional de Bioéthica para a Saúde (Ref: 498/CNBS/19); with further approval for a study modification (Ref: 149/CNBS/21). Deidentified quantitative data are available from the corresponding author on reasonable request, subject to PSI’s data sharing agreement.

## Results

From January 2020 to January 2023, 9,456 women were enrolled to the PEER project of whom 9,014 (95%) were eligible for hrHPV test (74 of the ineligible group were counseled for vFP only). Among all HPV-screened women, mean age was 37.7 years (sd =5.5). HIV positivity was 45.7%. Among the 60% who reported education level, 43% had primary and 29% had secondary schooling. Almost two thirds (64%, 5737/9014) were currently married (Table 1a). Uptake of integrated vFP services among those eligible for cervical cancer screening (or hrHPV test) was high. In total, 7,924 (88%) received both cervical cancer screening test and family planning counseling. Among those who received cervical cancer screening and vFP counseling, 1,694 (21%) took up modern method of vFP (initiated, switched, continued) on the day of counseling. Among those who took up a method of vFP, 20% (343/1694) reported never having used contraception previously (Figure 1).

**Figure 1:**
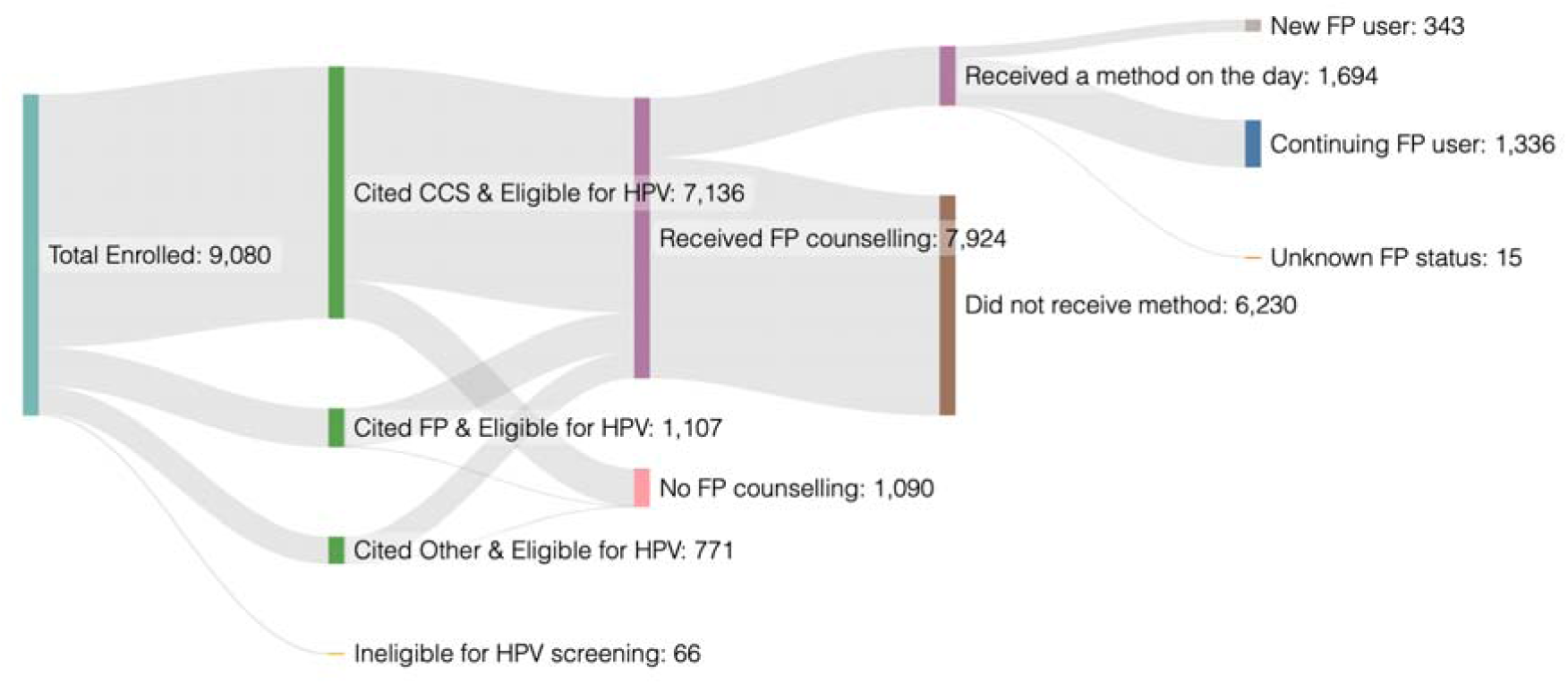
Sankey diagram of the integrated care pathway, showing total N enrolled excluding 66 participants ineligible for HPV screening. Disaggregation is by cited reason for attending clinic, and subsequent rates of flow through integrated vFP and method uptake.

**Table 1:** Characteristics of women enrolled in the PEER study, with disaggregation by receipt of integrated CCS and vFP services.

|  |  | **All women** | Integrated services received? |  | Significance |
|---|---|---|---|---|---|
|  |  |  | Yes | No |  |
| Total | N | **9014** | 7924 | 1090 |  |
|  | % | **100%** | 87.9% | 12.1% |  |
| Age (Years) | mean | **37.7** | 37.4 | 40.0 | *** |
|  | s.d. | **5.46** | 5.33 | 5.87 |  |
| Parity | mean | **3.16** | 3.27 | 2.37 | *** |
|  | s.d. | **1.8** | 1.78 | 1.74 |  |
| Province | Gaza | **47.4%** | 98.5% | 1.5% | *** |
|  | Maputo City | **52.8%** | 78.4% | 21.6% |  |
| Education | No formal education | **6.0%** | 6.4% | 2.5% | *** |
|  | Primary | **26.0%** | 26.5% | 21.8% |  |
|  | Middle school | **5.4%** | 4.9% | 9.0% |  |
|  | Secondary | **17.4%** | 17.3% | 18.4% |  |
|  | Technical/ higher | **5.6%** | 5.1% | 9.3% |  |
|  | Missing | **39.7%** | 39.8% | 39.0% |  |
| Marital status | Married/ in union | **63.7%** | 64.5% | 57.7% | *** |
|  | Separated/ divorced | **4.9%** | 5.3% | 2.1% |  |
|  | Single | **25.6%** | 24.4% | 34.3% |  |
|  | Widowed | **4.7%** | 4.6% | 5.1% |  |
|  | Missing | **1.2%** | 1.2% | 0.8% |  |
| HIV status | Negative | **54.0%** | 54.6% | 47.3% | *** |
|  | Positive | **46.0%** | 44.9% | 52.1% |  |
| Reason for consultation | HPV Screening | **79.2%** | 76.8% | 97.1% | *** |
|  | vFP | **12.3%** | 13.9% | 0.7% |  |
|  | Other | **8.4%** | 9.2% | 1.8% |  |
|  | Missing | **0.2%** | 0.1% | 0.4% |  |

\*\*\* = p value <0.001

Those who received integrated services were around 2.6 years younger, had on average around one child more, and were significantly less likely to report having HIV than those who did not receive integrated vFP and CCS (p>0.001) (Table 1). Those attending health services in Gaza province were significantly more likely to receive integrated services than those in Maputo City, and there appears to also be an education gradient, with those reporting lower levels of education being more likely to receive integrated care, although this variable has a lot of missing data. Those receiving integrated services were also more likely to be married.

A logistic regression model was fitted to estimate likelihood of receiving integrated CCS and vFP services (versus CCS alone) (Table 2). Education level was excluded from the model due to high levels of missingness. Multivariate analysis suggests that the likelihood of receiving integrated services is significantly associated with province, parity, age, marital and HIV status. Women in Maputo are significantly less likely to receive integrated care than those in Gaza (OR: 0.06, p<0.001), while being divorced/separated (compared to married), and having more children are associated with a greater likelihood of receiving integrated care (OR: 4.27, p<0.001 and OR: 1.30, p<0.001, respectively). Finally, being older, or reporting negative HIV status are both associated with a lower likelihood of receiving integrated care (OR: 0.90, p<0.001; and OR: 0.84, p<0.001, respectively).

**Table 2:** Results from logistic regression model examining the role of socioeconomic, demographic and HIV status variables on likelihood of receiving integrated services.

|  |  | **Odds ratio** | **95% Confidence intervals** |  | **p-value** |
|  |  |  | **Lower** | **Upper** |  |
|---|---|---|---|---|---|
| Province | Gaza | Ref |  |  |  |
|  | Maputo | **0.06** | 0.04 | 0.08 | <0.001 |
| Parity | (number of children) | **1.30** | 1.24 | 1.36 | <0.001 |
| Age | (years) | **0.90** | 0.88 | 0.91 | <0.001 |
| Marital status | Married/ in union | Ref |  |  |  |
|  | Separated/ divorced | **4.27** | 2.70 | 6.75 | <0.001 |
|  | Single | 0.93 | 0.79 | 1.08 | 0.33 |
|  | Widowed | **0.70** | 0.49 | 0.99 | 0.05 |
| HIV status | Negative | Ref |  |  |  |
|  | Positive | **0.84** | 0.73 | 0.97 | 0.02 |

N observations in model: 8,855
Pseudo R2: 0.226

Among the 7924 women who received integrated CCS and vFP counselling or services, 1694 reported receiving a method on the day of their visit, among whom 343 were new users, and 1336 were continuing users (figure 1). Among FP method users, the FP method mix was dominated by injectables and oral contraceptive pills (OCPs) (63.7% and 25.8%, respectively), with long-acting reversible contraceptives (LARCs) reported by around 1 in 20 women (3.6% and 1.0% for implants and IUDs, respectively), and condoms accounting for the remainder (5.8%).

In-depth interviews with clients and providers aimed to establish the degree to which they considered the integrated CCS&PT and vFP services to be acceptable and feasible, and to explore any barriers or enablers in their access to the program. In general, clients expressed positive opinions of the integration:

> *“I am very happy with the way the services are going, on the same day at the same place and with the same nurse, I save a lot of things at the same time, I save time because I only have one trip to the hospital, I save money on transport, before I used to get twice and now I get once and come here and thirdly I feel comfortable because I only talk to one nurse about my issues” (Client, Jose Macamo health facility, Maputo province)*

Another said:

> *“For me it [integration] is good the way it is because the users only have to expose their health problems to one health professional, they don’t have to join queues to receive two services, unlike what happened a few years ago” (Client, Chibuto health facility, Gaza province)*

Integrated care is highlighted as having multiple cost and time benefits for clients, as well as the implied benefit of additional privacy brought by only having to discuss health issues with one (rather than multiple) providers, sentiments shared by another interviewee:

> *“In the past you would go for screening and then you would go to another health unit … for family planning …nowadays I come here, and I do screening and vFP in the same place and the nurse doesn’t change. I feel more comfortable, because they are intimate matters that the fewer people know the better.” (Client, Jose Macamo health facility, Maputo city)*

For these clients, being seen by the same provider in a single location during the same visit not only made the service more convenient, but was also more psychologically comfortable, allowing a greater feeling of confidentiality around issues she considers sensitive. This theme emerged elsewhere:

> *“Let’s say I want to use the IUD planning method - I would have to undress in front of a person …and then have to go to another room to undress again for another person and it is a bit embarrassing. Now if it is to do both things with the same person for me it is more advantageous” (Client, Mavalane health facility, Maputo city)*

The perceived embarrassment of seeing health providers for SRHR-related services was reduced through having those services integrated into a single consultation. Providers also identified the improved privacy of a single visit as a key benefit of integration:

> *“[Clients] really fear removing clothes in front of people, so that is why they are more comfortable with this [integrated] service model” (Nurse, Jose Macamo health facility in Maputo city)*

Key informant interviews with clinic staff show that they also consider integrated services to be more convenient for their clients. One commented:

> *“The patients here have two services in one office so they come and they can do the vFP and the screening consecutively because it is also a constraint that [otherwise] she is in the family planning queue and after that queue is over, she comes to do the screening” (Provider, Jose Macamo health facility, Maputo city)*

In addition to convenience and privacy, some clients interviewed discussed how integration means learning about, and taking up services that they might not have been aware of otherwise:

> *“The mixed services have a greater effect than separate ones because many people who come for [family] planning end up being informed about screening and end up doing this and it is very good” (Client, Chibuto health facility, Gaza province)*

Some issues and barriers to accessing integrated services emerged through in-depth interviews, however:

> *“I think that they could take us who come for treatment out of the queue of people coming for screening and family planning for the first time, because we know where we are coming from and what we want while another group is coming for the first time and we are in the same queue, sometimes I feel pain because of sitting too much on those hospital benches.” (Client, Chicumbana health facility, Gaza province)*

Waiting times are an issue for this client, for whom the return visit for treatment involved having to wait alongside those being screened.

From the providers’ perspectives, the integration of vFP and CCS&PT services resulted in some unexpected outcomes:

> *“Because of the integration of services, I have noticed that most women have started to worry about their health, and some even bring their friends and even neighbors to the hospital for family planning and screening. One day we will have the pleasure of seeing all women coming to the hospital spontaneously for screening with self-sampling.” Provider, Mavalane health facility, Maputo city)*

Peer referral was not intended to be a component of the project, and yet the increased awareness of CCS&PT and vFP among women has led to word of mouth promotion in some communities. Meanwhile, the problems most discussed by providers concern the physical space and amenities available:

> *“We have problem in the offices for the work itself because that work needs privacy” (Provider, Chicumbana health facility, Gaza province)*

The importance of ensuring a private space for the provision of these integrated services cannot be overstated, particularly given the importance placed on privacy in clients’ narratives.

## Discussion

This study provides novel evidence of the uptake and acceptability of integrated CCS&PT and vFP services in Mozambique. Overall, we found high levels of uptake of integrated services among women enrolled in CCS&PT in the four public health facilities (in Maputo City and Gaza province), with almost 9 in 10 women who were enrolled for cervical cancer screening with hrHPV test also receiving vFP counselling on the day of their visit. Increased uptake of vFP when services are integrated has been observed elsewhere – including in Kenya, where HIV and vFP integration showed significant increases in FP method use (Chen et al., 2020). Around one in five women in this study who received these integrated services took up a contraceptive method on the day of their visit. These data suggest that offering vFP services to women who attend CCS&PT services in Mozambique is a feasible intervention, a finding supported by a recent systematic review (Bulstra et al., 2021). This presents an opportunity to improve access for women with unmet needs; one in five of the women taking up a contraceptive method were new users.

Clients who had accessed integrated services expressed a high degree of satisfaction with the convenience, enhanced privacy, reduced wait time and travel costs and opportunity to learn about new services that this model presents. Client satisfaction has been identified elsewhere to be an important driver of vFP service uptake (Paine et al., 2000) and a determinant of quality (Tessema et al., 2016). We found evidence of some sensitivity around sexual and reproductive health and rights service provision and exposure to providers among some interviewees. These understandable concerns were mitigated for many by an integrated model in which clients see a single provider for both vFP and CCS&PT. Further advantages, such as reduced time taken visiting health centers and reduced time waiting in queues within them were also important, particularly in the Mozambiquan context where transportation options to get to health centers for many clients may be limited, and the opportunity costs of multiple health center visits for those working in the informal economy, for example, may be considerable.

Providers who were involved in integrated service delivery expressed similar views to many clients concerning privacy and convenience. In addition, providers also discussed the impact that integrated services (and particularly clients having a positive experience) could have on increasing demand for these health services through peer referrals. The benefit of combining and making accessible vFP and CCS&PT for women in Mozambique has the potential to extend beyond a client, to her community and wider social network, including through word- of-mouth peer referrals.

Ensuring that services are truly integrated seems to be an important lesson from this research. Calls for sexual and reproductive health services to be brought fully and intentionally into universal health care (Appleford et al., 2020) in the literature support this conclusion. The key advantages stem from clients being able to see a single provider, in a single session, for both services. Other integration models involve multiple providers within the same clinic, and indeed one client discussed the inconvenience of having to queue for two separate sessions on the same visit. Future service provision should consider ensuring a client- centered, fully integrated model as the ideal approach.

Finally, while Mozambique’s national policy framework highlights the importance of providing integrated SRHR services, different models of integration exist, and practical implementation of an integrated model requires physical and administrative infrastructure that is not always prioritized in busy, clinical environments. Our study suggests that there may be efficiencies in full integrated services for both clients and providers, which might counter the upfront set-up effort.

## Limitations

This study has several limitations. First, the quantitative component relied on routine program data, with a challenge of missing data for some self-reported socio-demographic variables. Second, because this operational research was nested within a larger clinical trial, all participants in the study were recruited for CCS&PT; our definition of integration here therefore means the inclusion of vFP within a CCS&PT service provision model. We cannot comment therefore on the uptake or feasibility of a model where CCS&PT services are provided within an existing system of vFP service provision. Nevertheless, we did analyze clients’ self-reported original reason for clinic attendance, and a substantial minority reported attending for vFP, rather than CCS&PT. A final limitation regards the generalizability of the quantitative data, given the study population of those eligible for CCS&PT. Further research, outside of a clinical trial context would be useful to establish the degree to which our findings may apply to other groups of women in Mozambique.

## Conclusion

Integration of vFP into CCS&PT services resulted in increased overall vFP uptake, with a substantial minority of women taking up vFP for the first time through the integrated model. Our results suggest integration of these services is appealing to both women and providers, as it offers greater convenience, privacy, and fewer clinic visits. The implementation and potential scale-up of this model in Mozambique could represent a mutually beneficial opportunity to increase both access and uptake to voluntary family planning and cervical cancer screening and could result in increased efficiency in providing access to products and services that improve women’s sexual and reproductive health in ways that align with existing policy and are responsive to women’s needs.

## Data Availability

Deidentified quantitative data are available from the corresponding author on reasonable request subject to PSIs data sharing agreement.

## Acknowledgements

We are extremely grateful to the research participants, and health providers who generously gave of their time to share insights and data with the project. We would also like to express our appreciation to the data collection teams for their work.

*The PEER research group consists of:

The PEER Research Group*

Andrade, Viviane Andrade, ICON plc, São Paulo, Brazil

Andrea, Neves, Hospital Geral e Centro de Saúde José Macamo, Maputo, Mozambique Atif, Hira Atif, Ministério da Saúde de Moçambique (MISAU), Maputo, Mozambique Baker, Ellen Baker, Department of Gynecologic Oncology and Reproductive Medicine, The University of Texas MD Anderson Cancer Center, Houston, Texas, USA

Bouanchaud, Paul, PSI, London UK

Burny, Robert Burny, APOPO TB project, Maputo, Mozambique

Carla Carrilho, Carla, Universidade Eduardo Mondlane, Maputo, Mozambique

Carns, Jennifer Carns, Department of Bioengineering, Rice University, Houston, Texas, USA Castle, Philip E Castle, Department of Biostatistics, University of Texas MD Anderson Cancer Center, Houston, Texas, USA

Chissano, Marcos. PSI Mozambique, Maputo, Mozambique Chivambo, Edson PSI Mozambique, Maputo, Mozambique Chongo, Yolanda. PSI Mozambique, Maputo, Mozambique Dasa, Cristina Oliveira, Dasa, São Paulo, Brazil

Fellman, Bryan M Fellman,

Hoover, Hannah, PSI Mozambique, Maputo, Mozambique

Jean, Claude Nkundabatware, Hospital Provincial de Xai-Xai, Mozambique

Jeronimo, José Jeronimo, Department of Biostatistics, University of Texas MD Anderson Cancer Center, Houston, Texas, USA

Jessica Milan, Jessica, Department of Gynecologic Oncology and Reproductive Medicine, The University of Texas MD Anderson Cancer Center, Houston, Texas, USA

Kathleen M Schmeler, Kathleen M., Department of Gynecologic Oncology and Reproductive Medicine, The University of Texas MD Anderson Cancer Center, Houston, Texas, USA

Lathrop, Eva, PSI, Atlanta, USA

Lorenzoni, Cesaltina Lorenzoni, Ministry of Health, Maputo, Mozambique

Mariano, Arlete, A N Mariano, Hospital Geral e Centro de Saúde de Mavalane, Maputo, Mozambique

Mavume, Celda Mavume, Ministério da Saúde de Moçambique (MISAU), Maputo, Mozambique

Melissa Lopez Varon, Melissa

Mila, Pontremoli Salcedo, Department of Gynecologic Oncology and Reproductive Medicine, The University of Texas MD Anderson Cancer Center, Houston, Texas, USA

Monteiro, Eliane C S Monteiro, Hospital Geral e Centro de Saúde de Mavalane, Maputo, Mozambique

Mugolo, Rosita Paulo Mugolo, Ministério da Saúde de Moçambique (MISAU), Maputo, Mozambique

Nafissa, Osman, Universidade Eduardo Mondlane, Maputo, Mozambique

Rebecca Richards-Kortum, Rebecca, National Cancer Institute, Bethesda, Maryland, USA Ricardina Rangeiro, Ricardina, Hospital Central de Maputo, Maputo, Mozambique Thomas, Joseph P Thomas, Oncology Care & Research IS, University of Texas MD Anderson Cancer Center, Houston, Texas, USA

Tivir, Guilherima, PSI Mozambique, Maputo, Mozambique

